# How effective is vaccination protection?

**DOI:** 10.1101/2021.12.17.21267980

**Authors:** Milan Batista, Aleksej Turnšek

**Affiliations:** University of Ljubljana, Slovenia

## Abstract

This short note provides a simple static model to assess the effect of vaccination against hospitalization.

## 1 Introduction

The number of vaccinated admitted to hospitals can give us a false picture of the effectiveness of vaccination. Indeed, the proportion of hospitalized vaccinated is often expected to equal the proportion of the vaccinated in the population. Two limiting cases always hold: if no one is vaccinated, 100% of hospital admissions are unvaccinated; if everyone is vaccinated, 100% of hospital admissions are vaccinated. In general, however, estimating the fraction of hospitalized vaccinanted is not so straightforward.

In the following, a very simple static model will be given that can be used to assess the effectiveness of vaccination against hospitalization, i.e., a more severe course of the disease. As a measure of effectiveness, we choose the ratios between hospitalized unvaccinated to vaccinated in the population.

## 2 Model

Let *P* be the population size, *x* the fraction of vaccinated people, and 1 − *x* the fraction of unvaccinated. Let *H* be the cumulative number of hospitalized, *y* the fraction of vaccinated hospitalized, and 1 − *y* the fraction of hospitalized unvaccinated. We assume that the number of vaccinated hospitalized is proportional to the number of vaccinated people. Then we can write

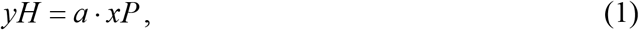

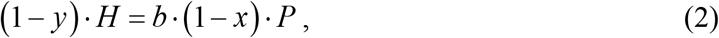

where *a* and *b* are the fractions of hospitalized vaccinated and unvaccinated in the population, respectively. The ratio between (2) and (1) is

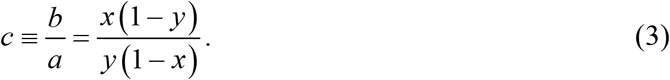

We take this ratio as a measure of vaccination effectiveness. Vaccination will be effective if *c* > 1, i.e., *b* > *a*, the chance of being hospitalized if unvaccinated is higher than if vaccinated. Note that*c* > 1imply *x* > *y*, i.e., vaccination is effective if the fraction of vaccinated in population is greater than the fraction of vaccinated in hospitals.

To determine *a* and *b*, we add (1) and (2), and introduce the fraction *α*, i.e., the fraction of people been hospitalized,

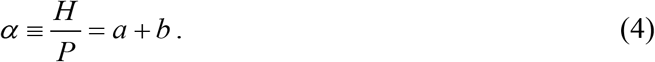

Taking into account (3), we get

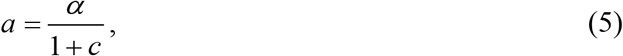

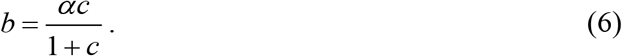

If we know *c* and assume it is a constant, then the fraction of hospitalized vaccinated equals

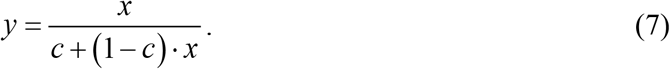

An example of a graph of this function is shown in Figure 1. The fraction of hospitalized vaccinated and unvaccinated will be the same when the proportion of vaccinated is *y* = 1/2. Now (7) yields

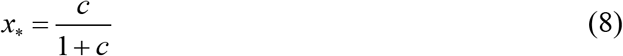

**Figure 1:**
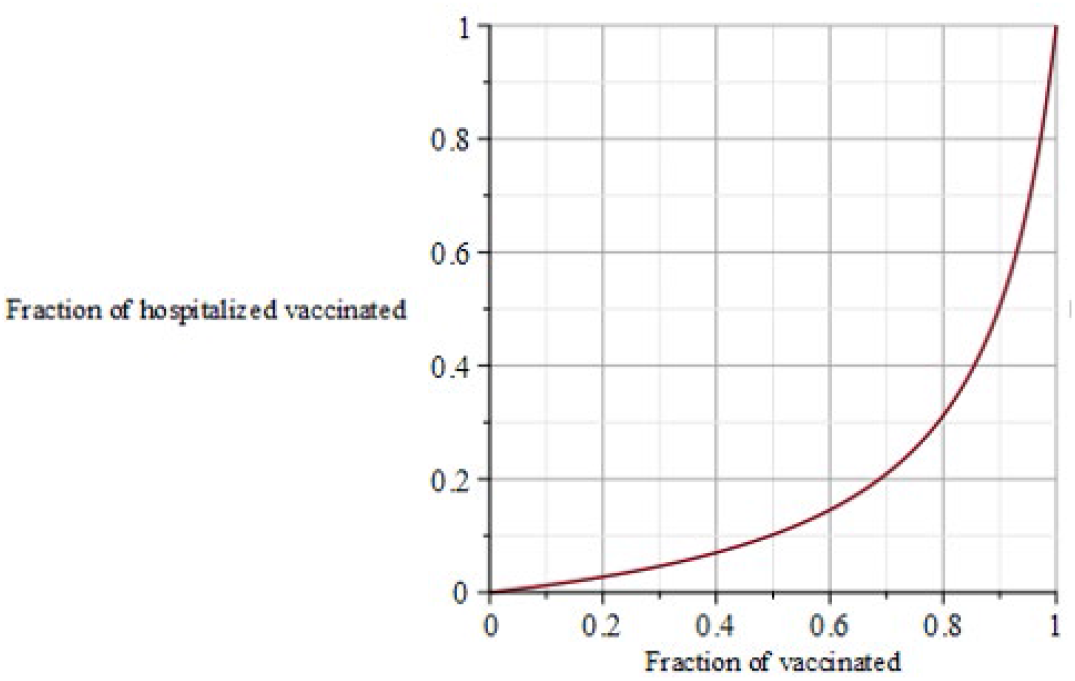
Graph of the proportion of hospital admissions according to the formula (8) for the case when *c* = 9

We note that the assumption for constant *c* is artificial and cannot generally hold. In fact, the vaccination effectiveness *c* is not constant, as can be seen from the graph in Figure 4, which shows the dependence of c on the proportion of fully vaccinated persons in the population for the course of the Covid-19 epidemic in Slovenia.

**Figure 2:**
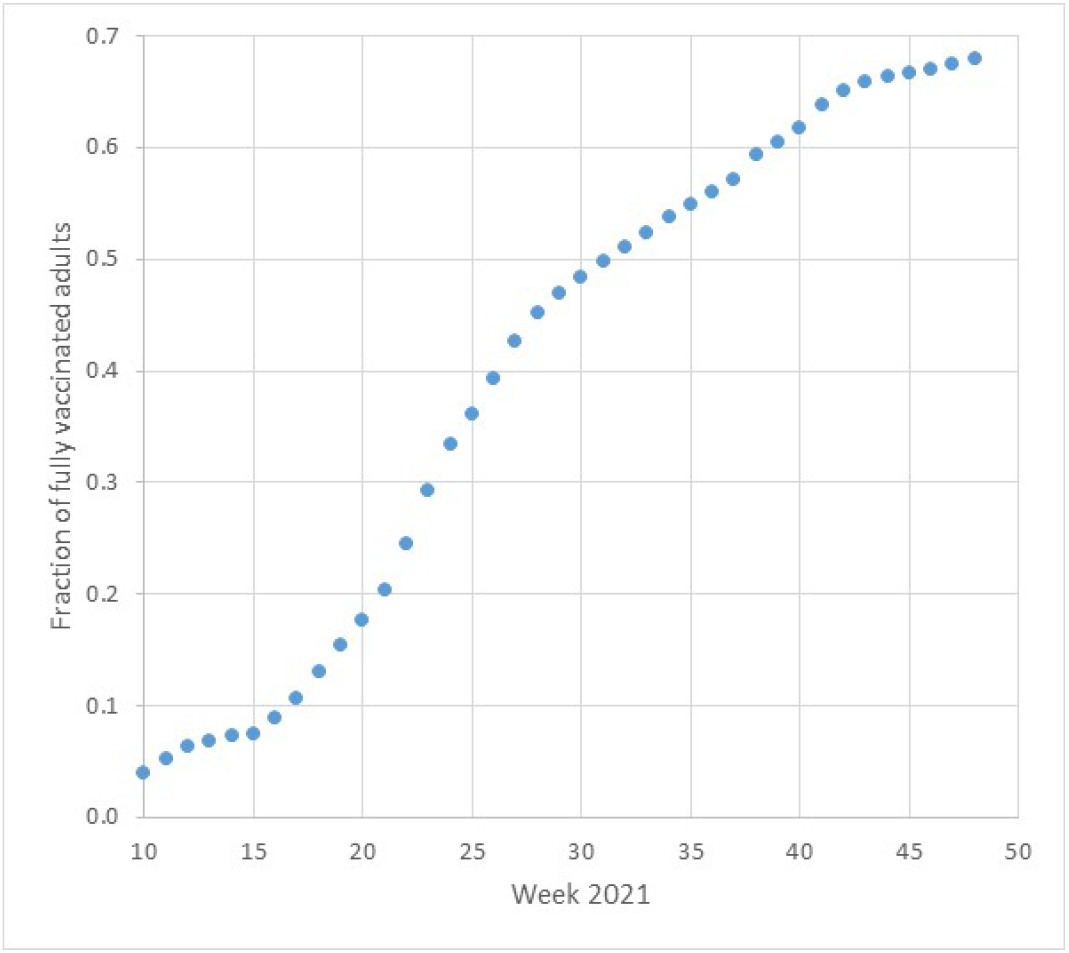
Fraction of fully vaccinated adults during the Covid-19 epidemic in Slovenia

**Figure 3:**
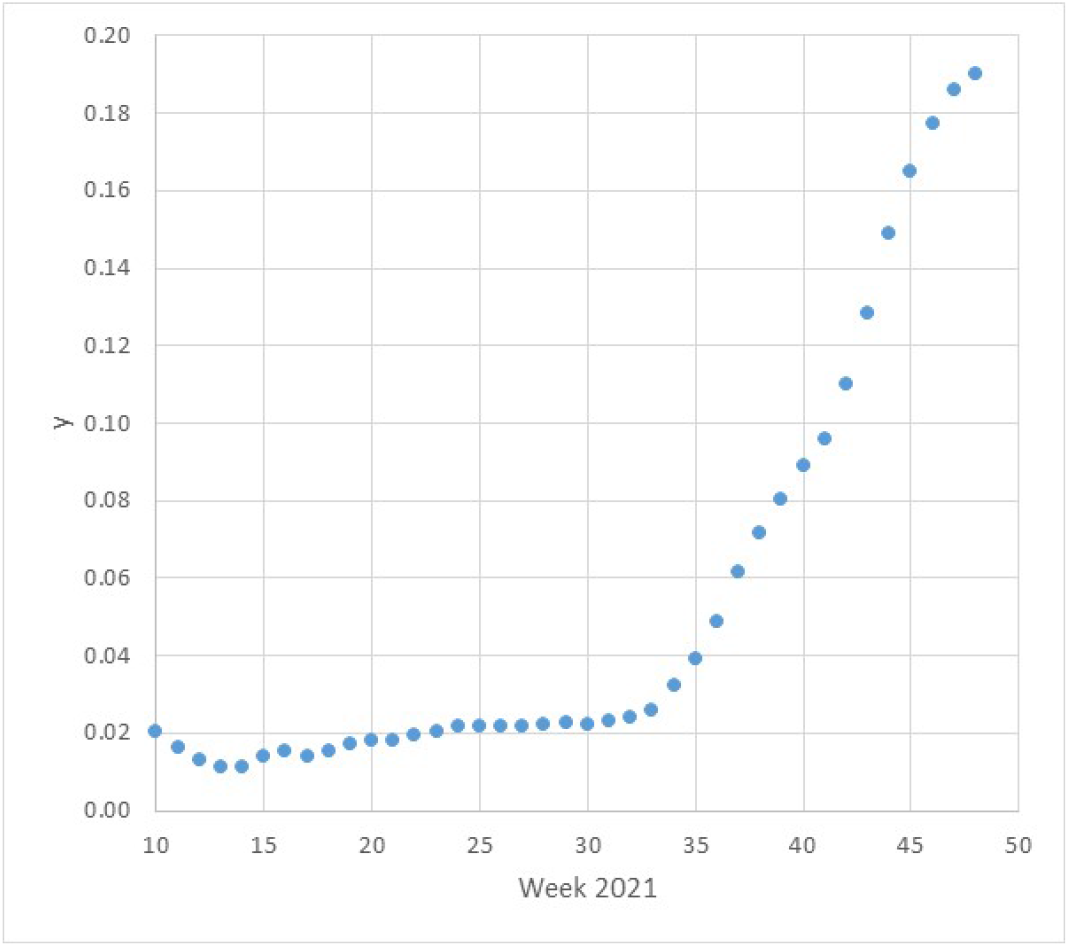
Fraction of hospital admissions with complete vaccination during the Covid-19 epidemic in Slovenia.

**Figure 4:**
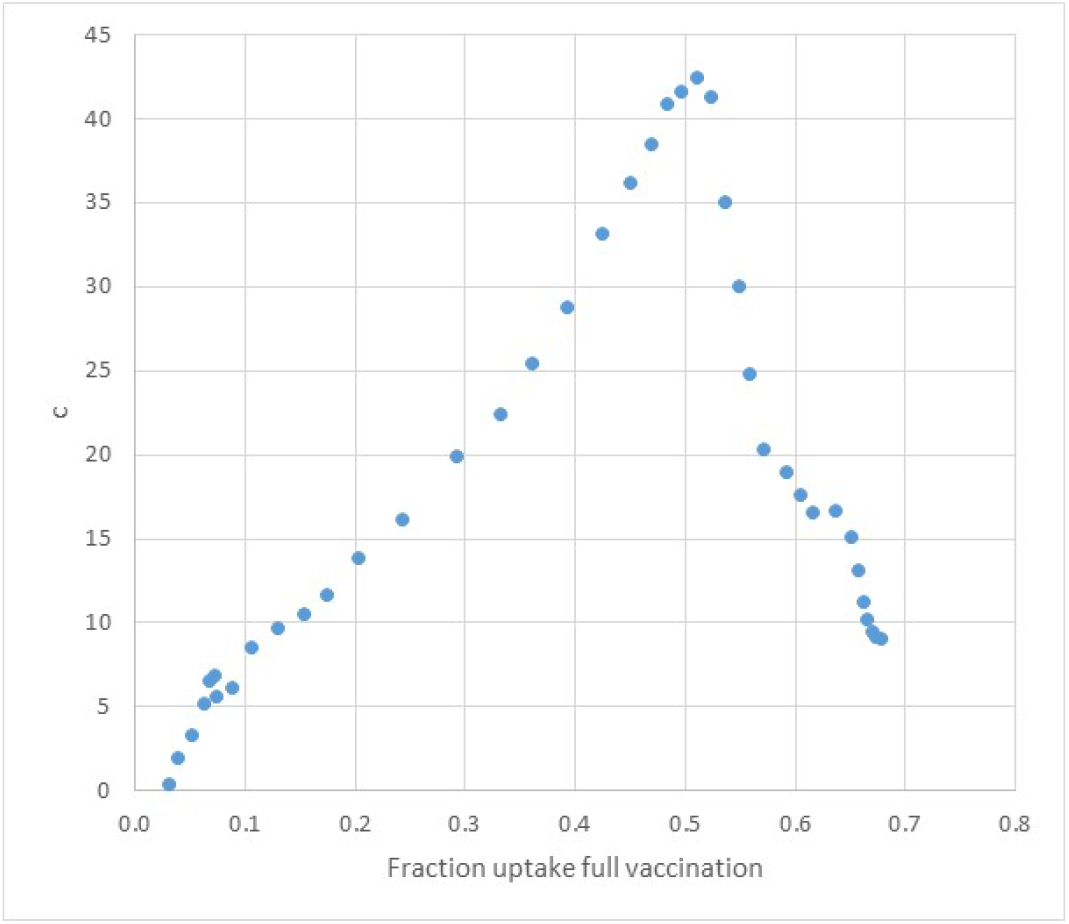
Calculated vaccination effectiveness *c* during the Covid-19 epidemic in Slovenia from March 2021 (week 9) to December 2021 (week 48).

## 3 Example

As a practical example, let us look at the calculation of vaccination effectiveness for the course of the Covid 19 epidemic in Slovenia in 2021. The data used are published on the Tracker website ^2^

Graphs 2 and 3 show the cumulative proportion of vaccinated adults in the population and among hospitalized patients.

Currently (Dec. 2021), 68% of the adult population in Slovenia is vaccinated, and the fraction of hospitalized vaccinated is currently 19%. Thus, for *x* = 0.68 and *y* = 0.19 we obtain, using (3),

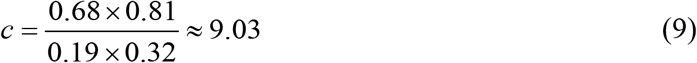

For the given data, this means that so far, the chance for unvaccinated to be hospitalized is about nine times greater than for vaccinated one.

If the above ratio were to be maintained, we would end up with a ratio of 1:1 in hospitals when the number of vaccinated

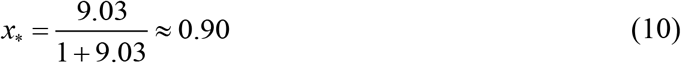

i.e., about 90%.

However, as already mentioned, vaccination effectiveness is not constant, as shown by the graph in Figure 4, which shows vaccination effectiveness against the proportion of fully vaccinated adults. In terms of time, the effectiveness of protection was highest somewhere around week 30, i.e., the beginning of August, when the proportion of vaccinated adults was about 50%, and the hospitals had only a few patients. With the new epidemic wave, the effectiveness started to drop, but in the last few weeks, it has stabilized at somewhere around 9.

In the Graph in Figure 5, the fractions of hospitalized nonvaccinated and vaccinated population is shown, where for calculation, we use Eq. (5) and (6). On the graph, one can easily observe that the population ratio reaches 9:1.

**Figure 5.**
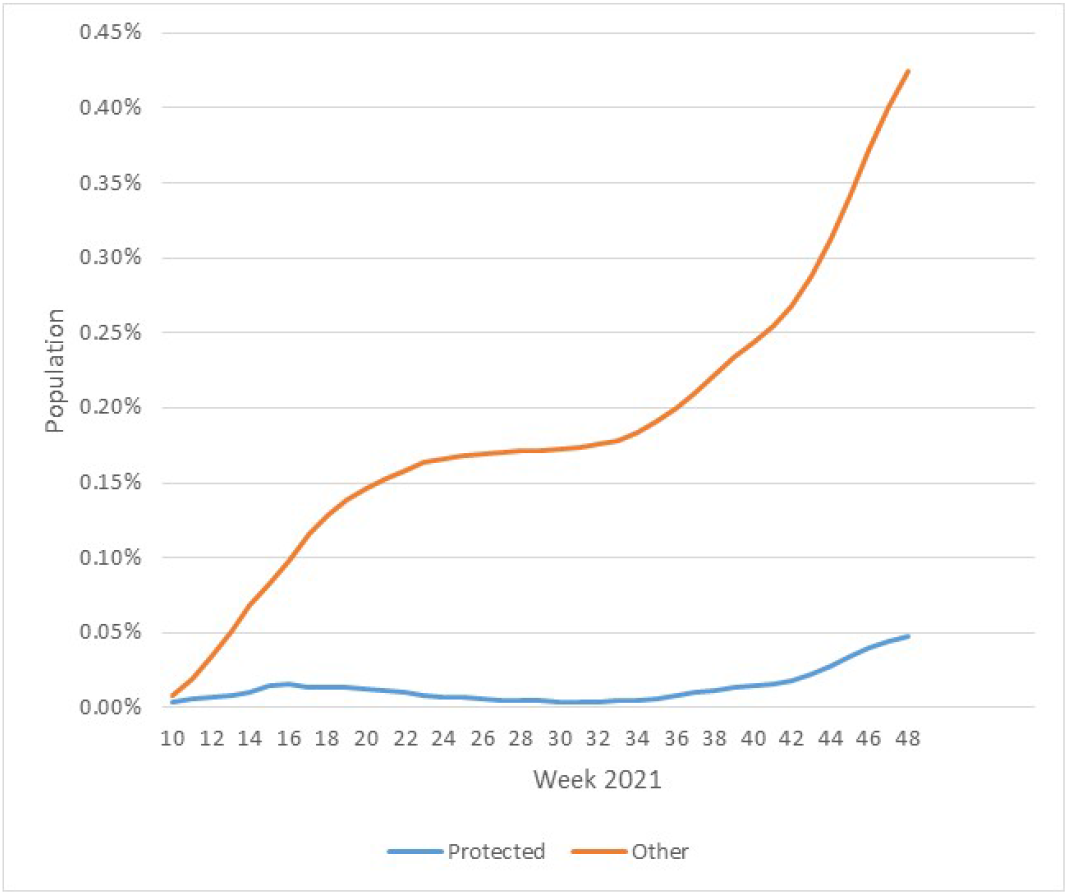
Fraction of people hospitalized for the Covid 19 epidemic in Slovenia.

## 4 Conclusion

This short paper provides a simple static model that can be used to assess the effectiveness of vaccination protection, i.e., the ratio between hospitalized vaccinated and unvaccinated in the population. The example shown is for hospitals, but it can easily be applied to the case of confirmed cases or victims of an epidemic.

## Data Availability

All data produced are available online at https://covid-19.sledilnik.org/en/stats

## Appendix

The model can be put into a probabilistic framework as follows. We have two events: event *V* that a person is vaccinated, and event *H* that a person was hospitalized. We use the bar over a letter to indicate that an event does not occur. Thus 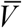 means that a person was not vaccinated, and 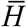 that person was not hospitalized.

We are interested in the conditional probability *P* (*H* |*V*) that a vaccinated person was hospitalized and the conditional probability 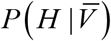 that the nonvaccinated person was hospitalized.

Using Bayes theorem, we have

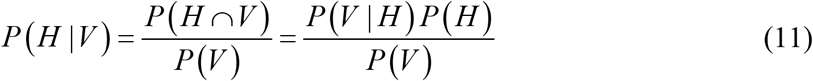

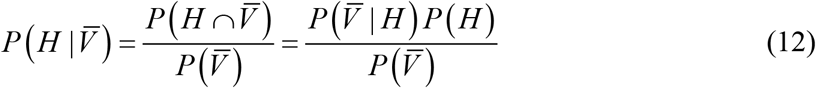

If the probability for hospitalization is *P* (*H*) = *α*, probability of being vaccinated is *P* (*V*) = *x*, and the probability that hospitalized is vaccinated is *P* (*V* | *H*) = *y* then the above equations can be rewritten into the following form:

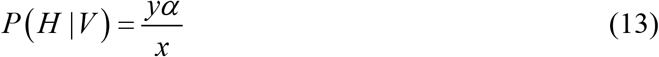

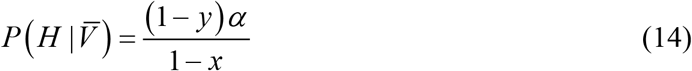

The ratio between these probabilities is

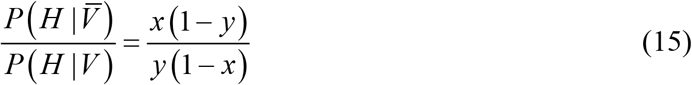

**Example**. So far (2021, week 48) in Slovenia was fully vaccinated 1146491 persons of population 2108977 (80%, i.e., 1687182 are adults), 7966 persons were hospitalized, and 1515 were vaccinated. We thus have

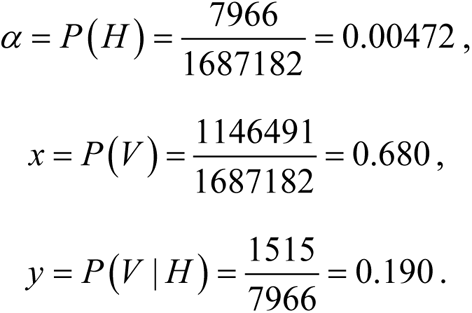

Using (4), (5), (6), we find

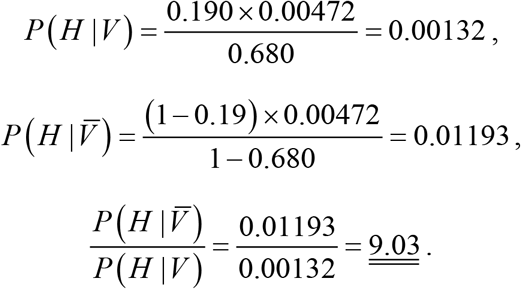

Thus the probability that an adult vaccinated was hospitalized is 0.1%, while the probability that a nonvaccinated adult was hospitalized is 1.2%. The ratio between the probabilities is 9:1.

## Footnotes

2 https://covid-19.sledilnik.org/en/stats

